# STRATEGY FOR THE CONTAINMENT, MITIGATION, AND SUPPRESSION OF THE COVID-19 PANDEMIC IN FRAGILIZED COMMUNITIES ON THE PERIPHERY OF A LARGE BRAZILIAN CITY

**DOI:** 10.1101/2020.09.28.20203174

**Authors:** V F Azevedo, LCB Peruffo, GM Nogueira, GKO Novakoski, FN Hajar, LK Rafael, RGO Brugnari, LE Vanzela, SB Larocca

## Abstract

**INTRODUCTION:** Prevention measures are highly important to poor communities because surveillance and access to health care may be limited.

**OBJECTIVES:** We aimed establish measures to contain and suppress the spread of COVID-19, associating education, active case tracking, and humanitarian aid in two needy communities in Brazil. The adherence to the measures and evolution of the number of cases were verified during the project.

**MATERIALS AND METHODS:** The target population consisted of approximately 1300 participants(350 families). A collection of epidemiological data was performed in family members registered for the project. Rapid tests were performed on people who had symptoms and their contacts. Scientific information through audio-visual materials,educational pamphlets written in colloquial language, food parcels,masks,hygiene and cleaning materials were provided directly to family nuclei.

**RESULTS:** The common needs faced by families were food inputs and/or ready-to-eat food, mentioned by 91.4% (233) of the people, and hygienic and cleaning materials, mentioned by 30.6% (78) of the people. Only 34.9% (84) of families had 70% rubbing alcohol or hand sanitizer gel at home.The most frequently cited sources of information on COVID-19 were television [cited by 82.4% (210) of the people]; social media [25.5% (65)]; friends, neighbours, or family members [13.7% (35)]; and radio [11.4% (29)] .A total of 83.7% (175) stated that the actions helped them to avoid leaving the community.

**CONCLUSIONS:** Community isolation may be the best way to contain the spread of pandemics in fragile populations with low socio-economic status.Educational actions combined with rapid testing and humanitarian aid were objective forms to promote community isolation.

## INTRODUCTION

The COVID-19 pandemic is an ongoing pandemic of an acute respiratory disease caused by the new severe acute respiratory syndrome coronavirus (SARS-CoV-2).^[1]^ The disease was first identified in Wuhan, Hubei Province, People’s Republic of China, on 1 December, 2019, and the first case was reported on 31 December of the same year.^[2,3]^ It is believed that the virus has a zoonotic origin because the first confirmed cases were mainly linked to the Huanan Wholesale Market, which also sold live animals.^[4-6]^ On 11 March 2020, the World Health Organization declared the outbreak a pandemic.^[7.8]^

In Brazil, the emergency alert was raised to level 2 (out of 3) on 28 January 2020, meaning it was considered an ‘imminent danger’ to the country.^[9]^ The notification of cases of COVID-19 is managed by the Integrated Health Surveillance Platform of Ministry of Health. We currently have had 3,501,975 cases and 112,304 deaths in Brazil.^[10]^

Prevention measures are highly important to poor communities because surveillance and access to health care may be limited.^[11]^ However, preventive measures such as frequent hand hygiene and social distancing are suboptimal in these populations. The lack of water and cleaning products hinders frequent hand washing and sanitizing of objects.^[12-14]^ A modelling study concluded that social distancing in a respiratory virus pandemic is 60 to 70% less effective in reducing the attack rate in an underdeveloped population than in a developed population.^[15]^ This lower effectiveness is attributed to greater numbers of people in the same household, which imply a higher proportion of intradomicile transmissions of the virus - which are not prevented by social distancing – out of the total transmissions.^[15]^ In addition, low-income individuals may be more averse to social distancing due to the need to work to provide food for their families, their lower flexibility in finding/changing jobs, their fear of losing their jobs, and their lack of formal jobs with working conditions set by law.^[12] [16,17]^ The lack of knowledge about the disease may also cause lower adherence to prevention measures. Knowledge about the disease is positively correlated with education and the adoption of prevention practices for both COVID-19^[18]^ and other diseases caused by respiratory viruses.^[19.20]^

There are several strategies to control an outbreak: containment, mitigation, and suppression. The containment measures are performed in the early stages of the outbreak and aim to locate and quarantine cases of infection, in addition to vaccination and other measures to control the infection to stop it spreading to the rest of the population. When it is no longer possible to contain the spread of the disease, the measures focus on delaying and mitigating its effects on society and the health system. Containment and mitigation measures can be performed simultaneously.^[21]^ Suppression measures require that more extreme measures be taken to reverse the pandemic by decreasing the reproductive number to less than 1.^[22]^

Part of the management of an outbreak of an infectious disease consists of trying to reduce the epidemiological peak, a process called ‘flattening the epidemiological curve’.^[23]^ This reduces the risk of overburdening health services and gives more time for new vaccines and treatments to be developed. Among the non-pharmacological interventions that control the outbreak are personal prevention measures, such as washing hands, wearing face masks, and voluntary quarantine; community prevention measures, such as closing schools and cancelling events that gather large numbers of people; environmental measures, such as cleaning and disinfecting surfaces; and measures that promote social adherence to these interventions.^[24]^ Among the suppression measures taken in some countries are quarantines of several cities, travel bans,^[25]^ mass screening, financial support for infected individuals so they isolate themselves, fines for those who break isolation, criminalization of stocking up on medical materials,^[26]^ and compulsory reporting of flu-like symptoms.

## OVERALL OBJECTIVES

This study aimed establish measures to contain and suppress the spread of COVID-19, associated with education, active case tracking, and humanitarian aid in two needy communities in the metropolitan region of Curitiba, Brazil, involving medical students from the Federal University of Paraná (UFPR) and volunteers. It also aimed to verify the effects of these measures on the outcome of adherence to the measures and evolution of the number of cases detected during the project. The general idea was to conduct a programme that could be globally reproduced and applied in any pandemic outbreak in fragile communities based on two basic and non-exclusive principles: education and humanitarian aid.

## MATERIALS AND METHODS

The target population of the present interventional study consisted of needy populations from the periphery of the municipalities of Curitiba (Caximba neighbourhood) and Araucária (Jardim Israelense neighbourhood), totalling 350 families (approximately 1,300 participants).

The Caximba community is located south of Curitiba. It has a population of predominantly European origin and an area of 8.22 km^2^.^[27]^ In 1989, a sanitary landfill was created in this region, which received waste from Curitiba and the metropolitan region.^[28]^ After the landfill stopped being used in mid-2009, ‘Vila 29 de Outubro’ was formed, the largest village in the region of the Caximba neighbourhood. Considered an irregular settlement area, since the land belongs to the Institute of Waters of Paraná, the community was built on flooded land, without basic sanitation.^[29]^ At least 1.1 thousand families inhabit these places unfit for dwelling.^[30]^

The Caximba neighbourhood has 767 households, with an average of 3.29 inhabitants per household. This makes this community more crowded than Curitiba, which has an average of 2.76 inhabitants per household.^[27]^ Thus, the site highlights the risk of spreading infectious diseases. In addition, only 4.44% of the households of this neighbourhood are connected to the general sewage network, raising the propensity to spread diseases that involve intestinal transmission.^[27]^

In the Capela Velha neighbourhood of Araucária, there are two large communities that were formed by land invasion: the Jardim Israelense community and 21 de Outubro community. Capela Velha is to the northwest Araucária and has approximately 25,000 inhabitants, 3.1% of whom are over 65 years old.^[31]^ The 21 de Outubro community was established in a portion of the Jardim Israelense affected by flooding near the Passaúna River dam. More than 300 families lived in this flooded portion until the Jardim Arvoredo II subdivision was completed, enabling approximately 170 families to be relocated.^[32]^ Data from the Department of Planning of the Araucária city hall show the poverty of the region: 60% of the residents have an income between one and two minimum wages and 13% of the residents an income below that amount.^[33]^

The patient care, educational procedures, and collection were done by students enrolled in health and medical courses of the medical schools of Curitiba and other volunteers.

The following procedures were performed:

1. Collection of epidemiological data from the studied populations as well as data on their knowledge of COVID-19
2. Performance of rapid tests on people from the community who had symptoms (suspicions) and their contacts. The tests were immunochromatography (intravenous blood collection and local verification of IgG/IgM positivity), which is indicated for patients with more than 10 days of symptoms suggestive of COVID-19, and RT-PCR, which is indicated for people with suspected active COVID-19 for less than 7 days.
3. Provision of scientific information and answering the population’s questions through audio-visual materials and educational pamphlets written in colloquial language, produced by the group of volunteers and distributed in the course of their activities.
4. Provision of food parcels, masks, hygiene, and cleaning materials directly to family nuclei to reduce the need for residents to go outside the community in search of humanitarian aid.

A questionnaire was applied in the form of a direct interview in the case of people with restricted or non-existent access to the Internet. For others, the questionnaire was published on the Google Forms platform, and its content was divided into three blocks:

a. Epidemiological data: age, sex, number of people living in the same household, educational level, income, presence of risk factors, etc.
b. Knowledge about the disease caused by SARS-CoV-2: main symptoms, measures for prevention and containment of virus transmission, where and when to seek medical care, etc.
c. Identification of symptomatic individuals (thus suspected of having COVID-19): current flu-like symptoms or symptoms within 30 days before the application of the questionnaire
d. The data collected on knowledge about COVID-19 and prevention measures were compared at 0 and 8 weeks after the beginning of the project.

## HUMANITARIAN AID

A web application was developed (https://voluntarioscontracovid19.web.app) to capture donations of money, food, and hygienic and cleaning materials; to publicize the project with photos; to recruit volunteers; and to post weekly updates on the activities of the team. The food or hygienic and cleaning materials that were donated in person were sorted and sanitized with hand sanitizer gel to prepare food parcels, which were distributed during the on-site activities in the communities. From March to August, the project captured more than 50 tons of food, hygienic materials, masks, clothes, toys, and household utensils, in addition to raising approximately $8,000 in cash donations. The donors learned about the project through social media and television.

## RESULTS

### EPIDEMIOLOGICAL DATA

A total of 255 people were included in the study. Each one represented a family registered for the project. A total of 79.6% (203) were women and 20.4% (52) were men, with a mean age of 39.9 (± 13.1) years [Table 1]. The mean number of people per family was 3.8 (± 1.6), the mean number of children or adolescents up to 16 years old was 1.6 (± 1.3) per family, and the mean number of elderly people per family was 0.2 (± 0.4). A total of 74.1% (189) of families had at least one person up to 16 years of age, and 14.1% (36) of families had at least one elderly person. A total of 8.6% (22) of the people were illiterate, 11.4%(29) had studied up to literacy, 48.2% (123) had studied through elementary school, 12.5% (32) had an incomplete secondary education, 15.7% (40) had a complete secondary education, 1.6% (4) had an incomplete higher education, and 0.4% (1) had a complete higher education.

**Tabel 1.**
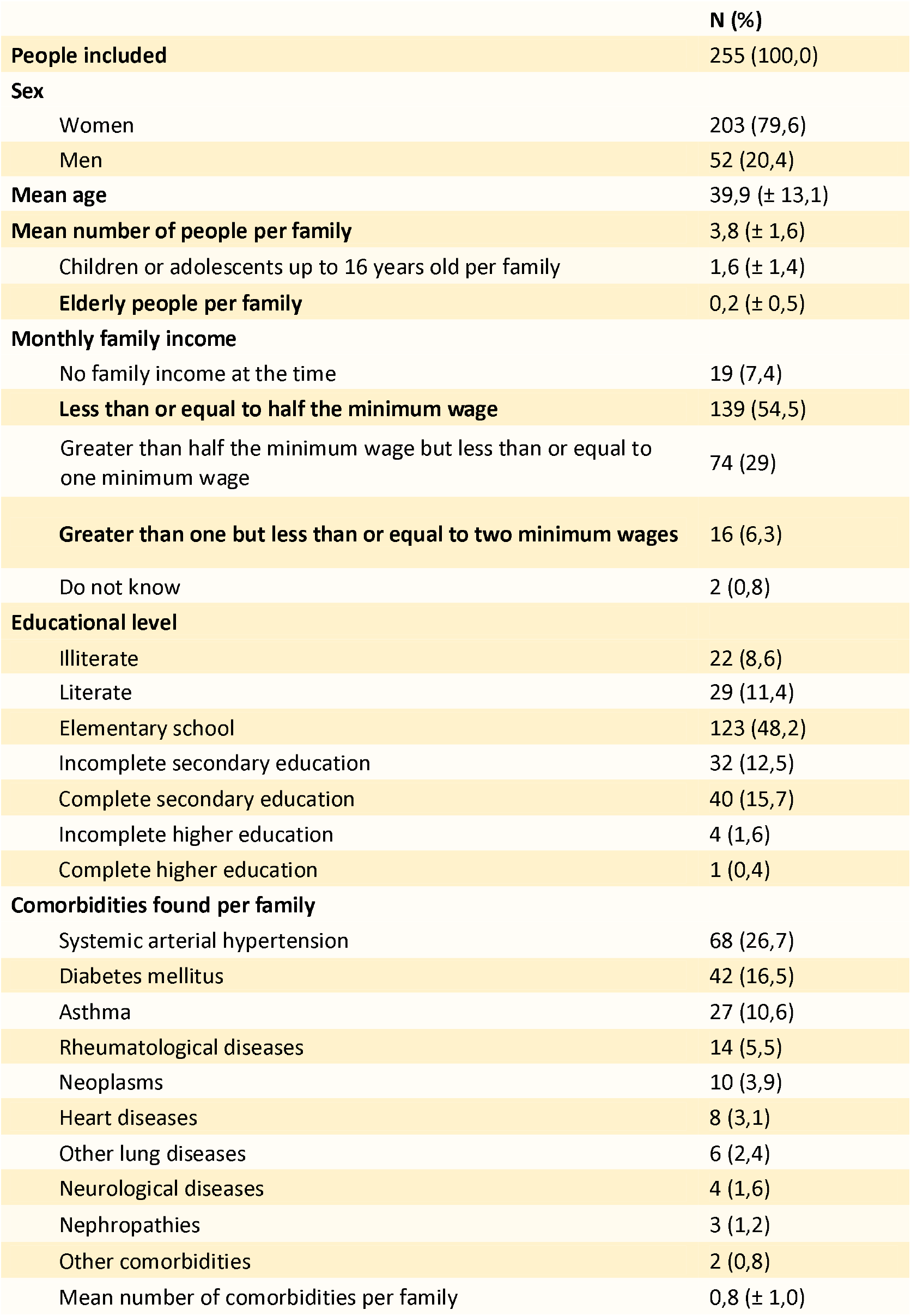
Socio Economic and Educational Data

|  | **N (%)** |
| --- | --- |
| **People included** | 255 (100,0) |
| **Sex** |  |
| Women | 203 (79,6) |
| Men | 52 (20,4) |
| **Mean age** | 39,9 (± 13,1) |
| **Mean number of people per family** | 3,8 (± 1,6) |
| Children or adolescents up to 16 years old per family | 1,6 (± 1,4) |
| **Elderly people per family** | 0,2 (± 0,5) |
| **Monthly family income** |  |
| No family income at the time | 19 (7,4) |
| **Less than or equal to half the minimum wage** | 139 (54,5) |
| Greater than half the minimum wage but less than or equal to one minimum wage | 74 (29) |
| **Greater than one but less than or equal to two minimum wages** | 16 (6,3) |
| Do not know | 2 (0,8) |
| **Educational level** |  |
| Illiterate | 22 (8,6) |
| Literate | 29 (11,4) |
| Elementary school | 123 (48,2) |
| Incomplete secondary education | 32 (12,5) |
| Complete secondary education | 40 (15,7) |
| Incomplete higher education | 4 (1,6) |
| Complete higher education | 1 (0,4) |
| **Comorbidities found per family** |  |
| Systemic arterial hypertension | 68 (26,7) |
| Diabetes mellitus | 42 (16,5) |
| Asthma | 27 (10,6) |
| Rheumatological diseases | 14 (5,5) |
| Neoplasms | 10 (3,9) |
| Heart diseases | 8 (3,1) |
| Other lung diseases | 6 (2,4) |
| Neurological diseases | 4 (1,6) |
| Nephropathies | 3 (1,2) |
| Other comorbidities | 2 (0,8) |
| Mean number of comorbidities per family | 0,8 (± 1,0) |

The monthly family income varied between zero and two minimum wages. A total of 7.4% (19) of the people reported not having a family income at the time, 54.5% (139) had a family income less than or equal to half the minimum wage, 29% (74) had a family income greater than half the minimum wage but less than or equal to one minimum wage, and 6.3% (16) had a family income greater than one but less than or equal to two minimum wages. The most common needs faced by families were food inputs and/or ready-to-eat food, a need that was mentioned by 91.4% (233) of the people, and hygienic and cleaning materials, mentioned by 30.6% (78) of the people. Only 34.9% (84) of families had 70% rubbing alcohol or hand sanitizer gel at home.

A total of 46.7% (119) the families had at least one person with comorbidities. The average number of comorbidities per family was 0.8 (± 1.0). The most frequent comorbidity was systemic arterial hypertension, which was present in 26.7% (68) of the families, followed by diabetes mellitus, which was present in 16.5% (42) of the families.

The most frequently cited sources of information on COVID-19 were television [cited by 82.4% (210) of the people]; social media [25.5% (65)]; friends, neighbours, or family members [13.7% (35)]; and radio [11.4% (29)] [Table 2]. A total of 25.5% (65) of the participants reported leaving the community regularly, 8.8% (22) doing so at least three times a week [Table 3]. Among the people who reported leaving the community, 43.9% (29) mentioned essential purchases – food and alcohol, for example – and 18.2% (12) mentioned work as a reason. A total of 33.3% (22) of the people who regularly left the community took public transportation. A total of 81.8% (206) answered that they could stay at home for 14 days in case of suspected or confirmed COVID-19.

**Table 2.** Sources of information on COVID-19

|  | **N (%)** |
| --- | --- |
| **Television** | 210 (82,4) |
| **Social media** | 65 (25,5) |
| **Friends, neighbours, or family members** | 35 (13,7) |
| **Radio** | 29 (11,4) |
| **Health workers** | 11 (4,3) |
| **Websites** | 8 (3,1) |
| **Government app** | 1 (0,4) |
| **Other sources of information** | 9 (3,5) |
| **No source of information** | 1 (0,4) |

**Table 3.** Reasons for leaving the community

|  | N (%) |
| --- | --- |
| **Essential purchases** | 29 (43,9) |
| **Work** | 12 (18,2) |
| **Medical care** | 12 (18,2) |
| **Bank** | 11 (16,7) |
| **Leisure** | 5 (7,6) |
| **Other reasons** | 6 (9,1) |

| Do you leave the community regularly? | N (%) |
| --- | --- |
| **Yes** | 65 (25,5) |
| Once a month | 4 (1,7) |
| Twice a month | 1 (0,4) |
| Once a week | 28 (11,1) |
| Twice a week | 10 (4,0) |
| Three times a week | 7 (2,8) |
| More than three times a week | 15 (6,0) |
| **No** | 187 (73,3) |

Do our actions help you avoid leaving the community?
|  | N (%) |
| --- | --- |
| **Yes, a lot** | 175 (83,7) |
| **Yes, a little** | 31 (14,8) |
| **No** | 3 (1,4) |

Only three people (1.4%) responded that our actions did not help them avoid leaving the community. A total of 83.7% (175) stated that the actions helped them a lot, and 14.8% (31) stated that the actions helped them a little to avoid leaving the community.

### COMMUNITY KNOWLEDGE ABOUT COVID-19

The mean number of COVID-19 symptoms correctly cited by the interviewees increased from 2.16 on the first test to 2.37 on the second test [Table 4]. The most cited were fever (from 66.3% to 72.5%), body pain (from 51.4% to 48.2%), dyspnoea (from 56.5% to 47.5%), and cough (from 40.4% to 43.5%). On the first test, 27.5% of people cited symptoms that are not commonly found in COVID-19, and 4.3% of people said they did not know any symptoms. In the second test, these percentages were 10.6% and 5.1%, respectively. A total of 47.5% of people at the time of the first test and 62.7% at the time of the second test knew that they would need to be quarantined for 14 days if they presented symptoms of COVID-19.

**Table 4.** COVID-19 symptoms

|  | First test | Second test | Difference | Confidence interval | p |
| --- | --- | --- | --- | --- | --- |
| Cough | 40,4 | 43,5 | 3,1 | -5,1; 11,4 | 0,493 |
| Fever | 66,3 | 72,5 | 6,3 | -1,5; 14,1 | 0,121 |
| Myalgia | 51,4 | 48,2 | -3,1 | -11,5; 5,2 | 0,501 |
| Difficulty breathing | **56,5** | **47,5** | **-9,0** | **-16,7; -1,4** | **0,021** |
| "I don't know" | 4,3 | 5,1 | 0,8 | -3,0; 4,6 | 0,824 |
| Wrong symptoms | **27,5** | **10,6** | **-16,9** | **-23,9; -9,9** | **<0,001** |
| Mean number of COVID-19 symptoms correctly cited by the interviewees | **2,2** | **2,4** | **0,2** | **0,0; 0,4** | **0,012** |
| Number of interviewees who correctly cited at least one COVID-19 symptom | 92,2 | 93,7 | 1,6 | -2,4; 5,6 | 0,523 |

The proportion of respondents who knew they did not need medical care in cases of mild symptoms of COVID-19 was 40.8% at the first test and 33.3% at the second test. In case of need, 36.7% of people at the first test and 58.8% at the second test would seek an emergency unit of the Brazilian Health System.

Regarding the forms of transmission of COVID-19, the most cited were close contact between people (69.0% on the first and 58.0% on the second test), saliva droplets (36.1%, 29.0%), and contaminated objects or surfaces (36.1%, 29.0%). The mean number of forms of transmission cited per person decreased from 1.65 to 1.46. The most frequently mentioned forms of prevention were washing hands (63.9% on the first and 63.1% on the second test), sanitizing the hands with alcohol (57.6%, 62.0%), wearing a mask when leaving the home (43.1%, 62.7%), staying in the house (51.8%, 46.7%), and social distancing (40.8%, 44.7%) [Table 5]. At the first test, 67.8 and 73.3% of respondents correctly answered what social distancing and isolation were, respectively. At the second test, these proportions were 71.8% and 78.8%.

**Table 5.** Forms of prevention

|  | First test | Second test | Difference | Confidence interval | p |
| --- | --- | --- | --- | --- | --- |
| Sanitizing the hands with alcohol | 57,6 | 62,0 | 4,3 | -3,8; 12,4 | 0,320 |
| Washing hands | 63,9 | 63,1 | -0,8 | -8,7; 7,1 | 0,919 |
| Wearing a mask when leaving the home | **43,1** | **62,7** | **19,6** | **11,3; 27,9** | **<0,001** |
| Social distancing | 40,8 | 44,7 | 3,9 | -4,2; 12,1 | 0,373 |
| Staying in the house | 51,8 | 46,7 | -5,1 | -13,6; 3,4 | 0,259 |
| Other correct forms of prevention | **2,4** | **0** | **-2,4** | **-4,6; 0** | **0,031** |
| "I don't know" | 2,4 | 2,0 | -0,4 | -3,3; 2,5 | 1 |
| Mean number of forms of prevention correctly cited by the interviewees | **2,6** | **2,8** | **-0,2** | **0,0; 0,4** | **0,049** |
| Number of interviewees who correctly cited at least one form of prevention | 97,3 | 98,0 | 0,8 | -2,0; 3,6 | 0,754 |

## DISCUSSION

Like transmission, mortality is higher in poor populations in a respiratory virus pandemic. In the 1918 and 1919 pandemics, mortality was seven times higher in poor regions, such as sub-Saharan Africa, Southeast Asia, and Latin America, than in developed regions, such as Europe and the United States.^[34,35]^ One study concluded that in a hypothetical pandemic similar to that of 1918, 96% of deaths would occur in developing countries.^[35]^ In addition, the mortality of an influenza pandemic is negatively correlated with per capita income, which alone explains approximately 50% of the variation in mortality.^[35]^ A 10% increase in per capita income implies a 9 to 10% reduction in mortality.^[35]^ This higher mortality of a respiratory virus pandemic in needy populations may be due, at least in part, to the prevalence of comorbidities such as tuberculosis, AIDS, and chronic malnutrition.^[16] [36]^ Factors discussed above, such as higher population density and poor housing conditions, may also contribute to increased mortality.^[37]^

Needy communities are especially vulnerable to pandemics. Although there are still no studies on the transmission of coronavirus in these communities, a study on a hypothetical pandemic of a new influenza strain estimated that the attack rate in underdeveloped populations may be 50% higher than the attack rate in developed populations, mainly due to the larger number of people living in the same household.^[15]^ Under conditions of overcrowded housing, small houses with poor ventilation, and housing with a single room, social distancing is impractical, as is the isolation of people who show symptoms, which predisposes the residents of the community to higher transmission of respiratory viruses.^[13] [38,39]^ Other factors that may contribute to the transmission of respiratory viruses among residents of poor communities are the sharing of utensils and the use of public transportation.^[16] [39]^

In addition to the disease itself, poor communities face difficulties indirectly caused by the pandemic, such as reduction or annulment of their income and loss of support for possible social actions that help the community.^[14]^ Thus, given the greater risks of COVID-19 transmission and mortality in needy communities, as well as the difficulties of implementing preventive measures and the aggravation of basic needs by the pandemic, the importance of actions specifically designed to help this population becomes clear, as the measures proposed for other populations are not efficient in these communities.

Although most of the people interviewed had no income or had an income less than one minimum wage, most had access to TV and social media, such as Facebook and Instagram, and obtained most of their information about COVID-19 from TV shows or national television news or from social media (Table 2). The application created by the Ministry of Health was rarely cited, and only one person accessed pandemic information through this public website. This demonstrates that public initiatives in Brazil need to be better publicized by the government to the poorest sections of the population.

As documented in the initial interviews and in the reapplication of the survey, most residents were already afraid to leave the community, and in general, they only did so for work reasons or to make purchases essential to their family’s subsistence. This facilitated our educational goal of emphasizing the need for residents to remain in the community during the pandemic.

While implementing this project and according to the data obtained from the registration of more than 300 families, we noticed that there was a potential circulation of the virus from the third week. Thus, we decided to apply a second questionnaire to assess the residents’ knowledge about COVID-19, as well as the reasons that led them to circumvent social distancing and expose themselves to the virus. In the fourth week, in addition to lunch boxes and food parcels, masks and bread were also distributed, which were obtained through partnerships with institutions that support movements and projects in fragile populations in Brazil. The second questionnaire was administered to residents already registered with the project.

Undoubtedly, populations with low education and economic development have difficulty understanding such a complex disease with so many different spectra as COVID-19. In addition, there was much confusing news with mixed messages in Brazil about measures of social distancing and isolation.^[40]^ It was encouraging to find that most respondents, as demonstrated by the retest, answered the questions with fewer errors. Undoubtedly, the fact that the residents received individual clarification on the modes of transmission of viral infection and its clinical manifestations, combined with the educational community actions through the production of short educational videos using colloquial language distributed via WhatsApp by our group, made a difference over the course of the project. This demonstrated that despite the benefit of community educational actions, individual educational actions are crucial for the adherence of residents of fragile populations to pandemic prevention measures.

A model in which community isolation in fragile populations is desired must include humanitarian aid activities. The provision of basic needs and help with hygienic and cleaning materials helps prevent residents leaving their community unnecessarily to seek basic supplies, exposing themselves to other infected communities.^[41,42]^ This is a pillar that we consider fundamental to the success of our project.

The COVID-19 pandemic in Brazil started with the upper-middle class, which had greater access to travel and contacts with foreign tourists in cities with large air hubs. Only then did the disease spread to needy and fragile communities.^[43]^ Although the Brazilian government establishes a monthly basic aid for up to two members of the same family (approximately US $120 monthly^[44]^), many did not have access to this amount because they needed to apply online, for which purpose a social security number would be necessary. Although we do not have the exact numbers, we found from community leaders in the two study areas that 10-20% of the children born there still did not have birth certificates. Brazil is a country of inequalities, with citizens who earn salaries comparable to those in a developed country and citizens who live in absolute poverty according to the World Health Organization definition. Thus, the actions it needs to take against a pandemic become complex and are not exhausted by public actions provided by the government at all levels. municipal, state, and federal. Rather, it must rely on the participation of members of organized civil society and non-governmental organizations for the success of the containment of pandemics. Brazilian public universities have a strong responsibility to enable aid programmes because they have trained professionals to solve the various problems that a fragile population may face in a pandemic such as COVID-19.

A potential shortcoming of our study was that we did not compare the impact of our actions on the spread of COVID-19 in other needy populations that did not have access to the same measures, near the locations where we operate. However, this was not our goal. Our goal was simply to know how much the population had improved in terms of their knowledge about the pandemic and how our activities improved their adherence to the containment measures.

## CONCLUSIONS

Community isolation may be the best way to contain the spread of pandemics in fragile populations with low socio-economic status. Educational actions combined with rapid testing and humanitarian aid are objective forms of aid that are well evaluated by these populations as promoting community isolation.

## Data Availability

All relevant data are included in the article, anyway authors can provide additional data once request with appropriate reason

**BIBLIOGRAPHY**

## REFERÊNCIAS BIBLIOGRÁFICAS

1. Organização Pan-americana da Saúde (OPAS). Folha informativa COVID-19: Escritório da OPAS e da OMS no Brasil [internet]. OPAS; 2020 [Acesso em: 01 abr. 2020]. Disponível em: https://www.paho.org/pt/covid19

2. Organização Mundial da Saúde (OMS). Coronavírus disease (Covid-19) pandemic [internet]. WHO; 2020 [Acesso em: 11 mar. 2020]. Disponível em: https://www.who.int/emergencies/diseases/novel-coronavirus-2019

3. Organização Mundial da Saúde (OMS). Statement on the second meeting of the International Health Regulations (2005) Emergency Committee regarding the outbreak of novel coronavirus (2019-nCoV) [internet]. WHO; 2020 [Acesso em: 23 jan. 2020]. Disponível em: https://www.who.int/news-room/detail/30-01-2020-statement-on-the-second-meeting-of-the-international-health-regulations-(2005)-emergency-committee-regarding-the-outbreak-of-novel-coronavirus-(2019-ncov)

4. Xu Wen Chen Qinhan. The Lancet disclosed the date of onset of the first patient with new coronavirus pneumonia, 7 days earlier than the oficial announcement (柳叶刀披露首例新冠肺炎患者发病日期,较官方通报早7天) [internet]. BJ News; 2020 [Acesso em: 01 fev. 2020]. Disponível em: http://www.bjnews.com.cn/news/2020/01/27/680493.html

5. Ma Danmeng. “The Lancet” published an article explaining the new coronavirus pneumonia, the first 41 cases showed signs of human transmission(《柳叶刀》 刊文详解新冠肺炎 最初41案例即有人传人迹象)[internet]. Caixin;2020[Acesso em 1 fev 2020]. 20 jan 2020. Disponível em: http://www.caixin.com/2020-01-26/101508497.html

6. Grupo de Epidemiologia do Mecanismo de Resposta a Emergências para Nova Pneumonia por Coronavírus do Centro Chinês de Controle e Prevenção de Doenças. Características epidemiológicas da nova pneumonia por Coronavírus (新型冠状病毒肺炎流行病学特征分析). Chinese Journal of Epidemiology, 2020. Disponível em: http://rs.yiigle.com/yufabiao/1181998.html

7. Moreira A, Pinheiro L. OMS declara pandemia de coronavírus. [internet]. G1; 2020 [Acesso em: 11 mar.2020].Disponível em: https://g1.globo.com/bemestar/coronavirus/noticia/2020/03/11/oms-declarapandemia-de-coronavirus.ghtml.

8. Branswell H, Joseph A. WHO declares the coronavirus outbreak a pandemic [internet]. STAT; 2020 [Acesso em: 11 mar. 2020] Disponível em: https://www.statnews.com/2020/03/11/who-declares-the-coronavirus-outbreakapandemic/#:~:text=The%20World%20Health%20Organization,said%20the%20situation%20will%20worsen.

9. Shinohara G. Coronavírus: Brasil sobe nível de alerta para ‘perigo iminente’ [internet]. O Globo; 2020 [Acesso em: 30 abr. 2020]. Disponível em: https://oglobo.globo.com/sociedade/coronavirus-brasil-sobe-nivel-de-alertapara-perigo-iminente-24215470

10. Plataforma Integrada de Vigilância em Saúde do Ministério da Saúde [homepage on the internet]. Painel de Monitoramento COVID-19. [accessed 20 Aug 2020]. Available in: http://plataforma.saude.gov.br/coronavirus/covid-19/

11. Bolton KJ, McCaw JM, Moss R, Morris RS, Wang S, Burma A, et al. Efficacité probable des interventions pharmaceutiques et non pharmaceutiques pour la réduction de la transmission du virus de la grippe en Mongolie. Bulletin de l’Organisation mondiale de la Santé. [periódicos da Internet] 2012 Abr [Acesso em 21 maio 2020]; 90(4): 264–71. Disponível em: https://www.who.int/bulletin/volumes/90/4/11-093419-ab/fr/

12. Bouye KE, Truman BI, Hutchins S, Richard R, Brown C, Guillory JA, et al. Pandemic influenza preparedness and response among public-housing residents, single-parent families, and low-income populations. Am J Public Health. [periódicos na Internet]. 2009 Out [Acesso em 21 maio 2020]; 99(2):287–93. Disponível em: https://pubmed.ncbi.nlm.nih.gov/19797740/

13. Rosenthal DM, Ucci M, Heys M, Hayward A, Lakhanpaul M. Impacts of COVID19 on vulnerable children in temporary accommodation in the UK. The Lancet Public Health [periódicos na Internet]. 2020 Mar [Acesso em 21 Mai 2020]; 5(5): 241–2. Disponível em http://dx.doi.org/10.1016/S2468-2667(20)30080-3

14. The Lancet. Redefining vulnerability in the era of COVID-19. The Lancet [editorial na Internet]. 2020 Abr [Acesso em 25 maio 2020] ;395 (10230):1089. Disponível em: http://dx.doi.org/10.1016/S0140-6736(20)30757-1

15. Milne GJ, Baskaran P, Halder N, Karl S, Kelso J. Pandemic influenza in Papua New Guinea: A modelling study comparison with pandemic spread in a developed country. BMJ Open [periódicos na Internet]. 2013 Mar [Acesso em: 10 mar. 2020]; 3 (3) Disponível em: https://bmjopen.bmj.com/content/bmjopen/3/3/e002518.full.pdf

16. Blumenshine P, Reingold A, Egerter S, Mockenhaupt R, Braveman P, Marks J. Pandemic influenza planning in the United States from a health disparities perspective. Emerg Infect Dis. [periódicos na internet] 2008 Mai [Acesso em 15 maio 2020]; 14(5): 709–15 Disponível em: https://pubmed.ncbi.nlm.nih.gov/18439350/.

17. Vaughan E, Tinker T. Effective health risk communication about pandemic influenza for vulnerable populations. Am J Public Health. [periódicos da Internet] 2009 Out [Acesso em 21 maio 2020]; 99(2):324–32 Disponível em https://pubmed.ncbi.nlm.nih.gov/19797744/.

18. Zhong BL, Luo W, Li HM, Zhang QQ, Liu XG, Li WT, et al. Knowledge, attitudes, and practices towards COVID-19 among chinese residents during the rapid rise period of the COVID-19 outbreak: A quick online cross-sectional survey. Int J Biol Sci. [periódicos na Internet]. 2020 Mar [Acesso em 15 abril 2020]; 16(10):1745–52. Disponível em: https://www.ncbi.nlm.nih.gov/pmc/articles/PMC7098034/

19. Maton T, Butraporn P, Kaewkangwal J, Fungladda W. Avian influenza protection knowledge, awareness, and behaviors in a high-risk population in Suphan Buri Province, Thailand. Southeast Asian J Trop Med Public Health. [periódicos na Intenret]. 2007 Mai [Acesso em 15 abril 2020] 38(3):560–8. Disponível em: https://pubmed.ncbi.nlm.nih.gov/17877234/

20. Paudel M, Acharya B, Adhikari M. Social determinants that lead to poor knowledge about, and inappropriate precautionary practices towards, avian influenza among butchers in Kathmandu, Nepal. Infect Dis Poverty. [periódicos na Internet] 2013 Jun [Acesso em 15 abril 2020] ; 2(1):1–10. Disponível em: https://www.ncbi.nlm.nih.gov/pmc/articles/PMC3710200/

21. Baird RP. What It Means to Contain and Mitigate the Coronavirus. [Internet]. The New Yorker; 2020 Mar [Acesso em 15 abril 2020]. Disponível em: https://www.newyorker.com/news/news-desk/what-it-means-to-contain-andmitigate-the-coronavirus

22. Ferguson NM, Laydon D, Nedjati-Gilani G, Imai N, Ainslie K, Baguelin M, et al. Impact of non-pharmaceutical interventions (NPIs) to reduce COVID-19 mortality and healthcare demand. Imperial College London. [periódicos na Internet] 2020 Mar; [Acesso em 15 maio 2020]; 1–20. Disponível em: https://spiral.imperial.ac.uk/bitstream/10044/1/77482/14/2020-03-16-COVID19-Report-9.pdf

23. Anderson RM, Heesterbeek H, Klinkenberg D, Hollingsworth TD. How will country-based mitigation measures influence the course of the COVID-19 epidemic? The Lancet [periódicos da Internet]. 2020 Mar [Acesso em 1 abril 2020]; 395(10228):931–4. Disponível em: https://pubmed.ncbi.nlm.nih.gov/32164834/

24. Qin A. China May Be Beating the Coronavirus, at a Painful Cost. [Internet] The New York Times; 7 mar 2020. [Acesso em: 1 abril 2020]. Disponível em: https://www.nytimes.com/2020/03/07/world/asia/china-coronavirus-cost.html

25. Cheung E. Wuhan pneumonia: Hong Kong widens net but can hospitals cope. [Internet]. South China Morning Post; Jan 2020 [Acesso em 1 abril 2020]. Disponível em: https://www.scmp.com/news/hong-kong/healthenvironment/article/3046634/wuhan-pneumonia-hong-kong-widens-netsuspected

26. AFP. Novo coronavírus pode ser transmitido pela fala, segundo cientistas dos EUA. [Internet]; IstoÉ; Abr 2020 [Acesso em 10 abril 2020]. Disponível em: https://istoe.com.br/novo-coronavirus-pode-ser-transmitido-pela-fala-segundocientistas-dos-eua/

27. Instituto de Pesquisa e Planejamento de Curitiba. Nosso Bairro/Caximba [base de dados da internet]. IPPUC: Curitiba, 2015. Acesso em: 18 abr 2020. Disponível em: https://ippuc.org.br/nossobairro/anexos/70-Caximba.pdf.

28. Secretaria Municipal do Meio Ambiente. Aterro Sanitário de Curitiba [Internet]. Prefeitura Municipal de Curitiba; 2020 [Acesso em 17 abril 2020]. Disponível em: https://www.curitiba.pr.gov.br/conteudo/aterro-sanitario-de-curitiba/454

29. Queiroz G. Caximba: Curitiba no limite [Internet]. Plural; Mar 2019 [acesso em 17 abr 2020]. Disponível em: https://www.plural.jor.br/noticias/vizinhanca/caximba-curitiba-no-limite/

30. Prefeitura Municipal de Curitiba. Comunidade tira dúvidas sobre o Bairro Novo da Caximba [Internet]. COHAB Curitiba; 2019 [acesso em 17 abr 2020]. Disponível em: https://www.curitiba.pr.gov.br/conteudo/aviso-lei-eleitoral/3174

31. População [homepage na Internet]. População Capela Velha -Araucária. Acesso em 17 abr 2020. Disponível em: http://populacao.net.br/populacao-capelavelha_araucaria_pr.html

32. Bem Paraná. Jardim Arvoredo começa a ser ocupado [Internet]. Bem Paraná; 2012. [acesso em 17 abr 2020]. Disponível em: https://www.bemparana.com.br/noticia/jardim-arvoredo-comeca-a-ser-ocupado238112#.XuJQMflKg2x

33. Barboza W. Regularização do 21 de outubro chega à última etapa [Internet]. O Popular; Jan 2019 [Acesso em 17 abr 2020]. Disponível em: https://www.opopularpr.com.br/regularizacao-do-21-de-outubro-chega-a-ultimaetapa/

34. Johnson NPAS, Mueller J. Updating the accounts: global mortality of the 1918-1920 “Spanish” influenza pandemic. Bull Hist Med [periódicos na Internet]. 2002 [Acesso em: 17 abril 2020]; 76(1):105–15. Disponível em: https://pubmed.ncbi.nlm.nih.gov/11875246/

35. Murray CJ, Lopez AD, Chin B, Feehan D, Hill KH. Estimation of potential global pandemic influenza mortality on the basis of vital registry data from the 1918-20 pandemic: a quantitative analysis. The Lancet. [periódicos na Internet]. Dez 2006 [Acesso em 17 abril 2020]; 368(9554): 2211–8. Disponível em: https://www.thelancet.com/journals/lancet/article/PIIS0140-6736(06)69895-4/fulltext

36. Archer B, Cohen C, Naidoo D, Thomas J, Makunga C, Blumberg L, Venter M, Timothy G, Puren A, McAnerney J, Cengimbo A, Schoub B. Interim report on pandemic H1N1 influenza virus infections in South Africa, April to October 2009: epidemiology and factors associated with fatal cases. Euro Surveillance [periódico na Internet]. Out 2009 [Acesso em 1 maio 2020]; 14(42) Disponível em: https://www.eurosurveillance.org/content/10.2807/ese.14.42.19369-en

37. Oshitani H, Kamigaki T, Suzuki A. Major issues and challenges of influenza pandemic preparedness in developing countries. Emerg Infect Dis. [periódicos na Internet]. Jun 2008 [Acesso em 1 maio 2020]; 14(6):875–80. Disponível em: https://wwwnc.cdc.gov/eid/article/14/6/07-0839_article

38. Bong CL, Brasher C, Chikumba E, McDougall R, Mellin-Olsen J, Enright A. The COVID-19 Pandemic: Effects on Low and Middle-Income Countries. Anesth Analg. [periódicos na internet]. 2020 Abr. [Acesso em 10 mar 2020]; 131(1):86-92. Disponível em: https://pubmed.ncbi.nlm.nih.gov/32243287/

39. Lima NNR, de Souza RI, Feitosa PWG, Moreira JL de S, da Silva CGL, Neto MLR. People experiencing homelessness: Their potential exposure to COVID-19. Psychiatry Res [periódicos na Internet]. 2020 Jun. [Acesso em 10 jul 2020]; 288 :112945. Disponível em: https://doi.org/10.1016/j.psychres.2020.112945

40. Jucá, B. “É inoportuno falar em flexibilizar isolamento quando vemos subir o número de mortos”. El País [Internet]. 2020 May 11 [cited 2020 Aug 20]. Available from: https://brasil.elpais.com/brasil/2020-05-12/e-inoportuno-falar-em-flexibilizar-isolamento-quando-vemos-subir-o-numero-de-mortos.html

41. Organização Mundial da Saúde (OMS). Covid-19 Strategy Update [internet]. WHO; Abr 2020 [Acesso em: 21 maio 2020]. Disponível em: https://www.who.int/publications/i/item/covid-19-strategy-update14-april-2020

42. Santos JAF. Covid-19, causas fundamentais,classe social e território. Trab Educ e Saúde. 2020;18(3).

43. de Souza WM, Buss LF, Candido D da S, Carrera JP, Li S, Zarebski AE, et al. Epidemiological and clinical characteristics of the COVID-19 epidemic in Brazil. Nat Hum Behav. 2020;1–19.

44. Governo Federal - Ministério da Cidadania [homepage on the internet]. Auxílio Emergencial [accessed 20 Aug 2020]. Available in: https://www.gov.br/cidadania/pt-br/servicos/auxilio-emergencial

